# Resilience and resistance to Alzheimer’s disease-associated neuropathological substrates in centenarians: an age-continuous perspective

**DOI:** 10.1101/2022.08.28.22279304

**Authors:** Meng Zhang, Andrea B. Ganz, Susan Rohde, Annemieke J. M. Rozemuller, Netherlands Brain Bank, Marcel J.T. Reinders, Philip Scheltens, Marc Hulsman, Jeroen J.M. Hoozemans, Henne Holstege

## Abstract

**INTRODUCTION:** With increasing age, neuropathology associated with Alzheimer’s disease (AD) accumulates in brains of cognitively healthy individuals: are they resilient or resistant against AD-associated neuropathologies?

**METHODS:** In 85 centenarian brains, we correlated NIA-Amyloid stages, Braak-NFT stages and CERAD-NP scores with cognitive performance determined close to death (MMSE). We assessed centenarian brains in context of 2,131 brains from AD patients, non-AD demented and non-demented individuals (age-range 16-100+ years).

**RESULTS:** With age, brains from non-demented individuals reached NIA-Amyloid and Braak-NFT stages as observed in AD patients, while CERAD-NP scores remained lower. In centenarians, NIA-Amyloid stages varied (22.4% had the highest stage 3), Braak-NFT stages rarely exceeded IV (5.9% had stage V), and CERAD-NP scores rarely exceeded 2 (4.7% had score 3); within these distributions, we observed no correlation with MMSE (NIA-Amyloid: *P*=0.60; Braak-NFT: *P*=0.08; CERAD-NP: *P*=0.16).

**DISCUSSION:** Cognitive health can be maintained despite the accumulation of high levels of AD-related neuropathological substrates.

## 1 Background

Alzheimer’s disease (AD) is associated with the extracellular accumulation of (1) amyloid beta (Aβ) plaques or (2) neuritic plaques (NPs) and (3) the intracellular aggregation of phosphorylated tau protein into neurofibrillary tangles (NFTs).[1] Previous studies have indicated that the brains of middle-aged AD patients have accumulated the highest levels of most AD-associated neuropathological hallmarks, while the brains of cognitively healthy middle-aged individuals are almost free of these neuropathological substrates.[2,3] Intriguingly, after middle age, the levels of these neuropathological substrates were observed to increase with age in the post-mortem brains of cognitively healthy individuals.[2–5] In fact, a large autopsy study found that 30-40% of the brains from 79-year-olds harbor significant AD-associated neuropathological changes, while only 15% of these elderly were clinically diagnosed with AD.[6,7] In line with this, we and others previously observed that the levels of these neuropathological substrates are highly variable in nonagenarians and centenarians with diverse cognitive performance.[4,8–10] This variability in the level of cognitive performance and the level of neuropathological substrates represents a window of opportunity to investigate to what extent the different neuropathological substrates associate with cognitive performance at high ages. Further, this allows to investigate whether maintained cognitive performance during aging depends on being tolerant to the effects of accumulated AD-associated neuropathological substrates (resilience) or whether it depends on avoiding the build-up of AD-associated neuropathological substrates (resistance).

Here, we correlated the levels of neuropathological substrates observed in the brains of 85 centenarians with cognitive performance determined close to death by the Mini Mental State Examination (MMSE). Next, we compared these levels of AD-associated neuropathological substrates with those observed in the brains of 2,131 individuals, representing an age-continuum from 16 to 100+ years (851 AD cases, 654 non-demented controls, and 626 non-AD demented individuals). This allowed us to determine (1) to what extent the levels of neuropathological substrates change with age; (2) the effect of age on the potential of each AD-associated pathological substrate to distinguish between AD and cognitive health; and (3) how the inter-correlation between the levels of different pathological substrates changes with increasing age.

## 2 Methods

### 2.1 100-plus Study cohort

We included brains donated by 85 centenarians (ages at death: 100-111) who died between 2013 and 2021 and self-reported to be cognitively healthy at inclusion in the 100-plus Study cohort [11], confirmed by a proxy. Global cognitive performance was assessed during a baseline visit and yearly follow-up visits, and included the MMSE, an 11-item test with a maximum score of 30 points.[8,11] Scores were imputed for missing values when <6 of the 30 points could not be scored due to sensory deficits such as hearing and vision impairment and/or general fatigue [8], otherwise MMSE was set to “missing”.

### 2.2 Netherlands Brain Bank (NBB) cohort

Neuropathology data was obtained from 2,131 individuals, AD cases (AD), non-demented individuals (ND) or non-AD-demented individuals (non-AD), who agreed to brain donation to the Netherlands Brain Bank (NBB, www.brainbank.nl) between 1979 and 2018.

### 2.3 Neuropathological assessment

Autopsies and neuropathological assessments for the NBB-cohort and the 100-plus Study cohort were performed by the NBB, as described in the Supplementary Material. For all donated brains, we evaluated the level or spread of four AD-associated neuropathological substrates: (1) NIA Amyloid stage, (2) Braak-NFT stage, (3) CERAD-NP score, and (4) sex-corrected brain weight.[12] See Supplementary Material for details.

### 2.4 AD vs ND comparison across age-continuum

To assess the age-related changes in the levels of AD-associated neuropathological substrates, we applied a dynamic 25-point sliding window across the ages of AD cases and ND individuals from the NBB separately. In this method, the neuropathology levels in the AD and ND brains were sorted according to age-at-death. For each neuropathological substrate, the mean level of each 25-point window was calculated. Each window encompassed a set of 12 cases with ages lower and 12 cases with ages higher than the age of the central case. Across each window-set, we calculated a confidence interval (CI) with 5% increments to indicate the distribution of pathology levels (i.e. 5%, 10%, …, 90%, 95% CI). Next, for each sliding window position, we calculated the difference in the average neuropathological levels between AD cases and ND individuals. The CI of the difference with 5% increments was determined by bootstrapping (n=1000).

### 2.5 Distribution of neuropathology levels in age interval

The diverse levels of neuropathological substrates and cognitive performance of centenarians allow them to be diagnosed as either AD, non-dementia, or non-AD dementia. Thus, to fairly compare the distributions of neuropathological levels between the centenarian cohort and the NBB cohort, we added the non-AD demented individuals from the NBB cohort to the distribution. The distributions of the level of each neuropathological substrate in AD cases, ND controls, and non-AD individuals were estimated separately for each age interval (i.e., ≤60, 60-70, 70-80, 80-90, ≥90) and visualized by generating density plots using a Gaussian kernel. Next, the overall distribution of the level of each neuropathological substrate in the NBB cohort for each age interval was estimated by summation of the densities of AD cases, ND controls, and non-AD individuals across neuropathological levels.

### 2.6 Pairwise correlation between different neuropathology levels

To evaluate the pairwise correlations between (1) NIA Amyloid stage and Braak-NFT stage, (2) NIA Amyloid stage and CERAD-NP score, and (3) Braak-NFT stage and CERAD-NP score with age, we merged the AD cases, non-demented controls and non-AD demented individuals as one cohort. We used a 51-point sliding window, which was constructed in the same way as the 25-point window, but using 25 cases with ages lower and 25 cases with ages higher than the age of the central brain sample. For each neuropathology pair and sliding window position, we calculated the Pearson correlation coefficient and corresponding CIs (5% increments).

### 2.7 Statistical analyses

We applied a linear regression model to test the association between each neuropathological substrate and MMSE in the centenarian cohort. All regressions were corrected for sex, education, and time between last acquired MMSE and death. All calculations were performed using R (version 3.6.3). Pearson correlation, and linear regression were performed using the “stats” R package.[13]

## 3 Results

### 3.1 Sample characteristics

The demographic characteristics (age, sex, education), cognitive performance, and neuropathological assessments of 85 centenarian brain donors are shown in Table 1. At study inclusion, which occurred a median of 25 months before brain donation (IQR: 13-36), the median MMSE score across the 85 centenarians (74% female) was 26 (IQR: 23-28). At the last study visit available, a median of 9 months prior to brain donation (IQR: 4-13), the median MMSE score was 25 (IQR: 22-27).

**Table 1:** Characteristics of the centenarians in the 100-plus Study cohort

|  | 100-plus Study cohort |  |
| --- | --- | --- |
|  | n | median (IQR) |
| Clinical demographics |  |  |
| Age [y] | 85 | 103.2 (102.3-104.6) |
| Female/male | 63/22 | — |
| Education | 85 | 3 (1-4) |
| MMSE | 82 | 25 (22-27) |
| Neuropathological substrates |  |  |
| NIA amyloid stage | 85 | 2 (1-2); 0: 9.4%, 1: 35.3%, 2: 32.9%, 3: 22.4% |
| Braak-NFT stage | 85 | 3 (3-4); I: 2.4%, II: 14.1%, III: 42.4%, IV: 35.3%, V: 5.9% |
| CERAD-NP score | 85 | 1 (0-2); 0: 43.5%, 1: 29.4%, 2: 22.4%, 3: 4.7% |
| Brain weight [gr] | F: 63; M: 22 | F: 1,067 (1,005-1,125); M: 1,165 (1,091-1,220)<br>Sex-corrected: 1,068 (1,005-1,125) |

Based on clinical data that was provided upon autopsy and observed post-mortem neuropathology, the 2,131 NBB brain donors (56% female) were diagnosed as AD cases (AD; n=851, aged 37-102), non-demented controls (ND; n=654, aged 16-103), or non-AD demented individuals (non-AD; n=626, aged 16-103) (Table 2). Non-AD dementia individuals died with or from diverse dementia subtypes and age-related pathology: Fronto-temporal dementia (FTP, 35.3%), Primary age-related tauopathy (PART, 26.0%), Parkinson’s disease (18.4%), Vascular dementia (15.2%), or other (5.1%), see Supplementary Table S1.

**Table 2:** Characteristics of the NBB donors according to age at death

| Characteristic | <60 Yr | 60-70 Yr | 70-80 Yr | 80-90 Yr | ≥90 Yr | Total |
| --- | --- | --- | --- | --- | --- | --- |
|  | N, median<br>(IQR) | N, median<br>(IQR) | N, median<br>(IQR) | N, median<br>(IQR) | N, median<br>(IQR) | N, median<br>(IQR) |
| Female/Male (N) | 103/168 | 158/202 | 268/284 | 450/231 | 210/57 | 1,189/942 |
| Neuropathological diagnosis |  |  |  |  |  |  |
| **Alzheimer's disease (N)** | **42** | **132** | **204** | **323** | **150** | **851** |
| NIA Amyloid stage | 29, 3 (3-3) | 75, 3 (3-3) | 120, 3 (3-3) | 159, 3 (3-3) | 77, 3 (3-3) | 460, 3 (3-3) |
| Braak-NFT stage | 36, VI (VI-VI) | 110, VI (V-VI) | 178, V (V-VI) | 292, V (V-VI) | 137, V (IV-V) | 753, V (V-VI) |
| CERAD-NP score | 5, 3 (3-3) | 16, 3 (3-3) | 29, 3 (3-3) | 28, 3 (2-3) | 17, 3 (2-3) | 95, 3 (3-3) |
| Brain weight [gr] | 42, 1,017<br>(950-1,103) | 132, 985<br>(890-1,065) | 204, 995 (913-1,088) | 323, 1,010<br>(941-1,084) | 150, 959<br>(917-1,052) | 851, 998 (920-1,078) |
| **Non-dementia (N)** | **180** | **90** | **161** | **158** | **65** | **654** |
| NIA Amyloid stage | 18, 0 (0-0) | 17, 0 (0-1) | 65, 1 (1-2) | 90, 1 (0-2) | 47, 2 (1-2) | 237, 1 (0-2) |
| Braak-NFT stage | 52, 0 (0-0) | 45, I (0-I) | 113, I (I-II) | 129, II (I-II) | 55, II (I-III) | 394, I (0-II) |
| CERAD-NP score | 16, 0 (0-0) | 11, 0 (0-0) | 27, 0 (0-0) | 36, 0 (0-0) | 23, 0 (0-1) | 113, 0 (0-0) |
| Brain weight [gr] | 180, 1,267<br>(1,190-1,360) | 90, 1,225<br>(1,130-1,274) | 161, 1,160<br>(1,077-1,232) | 158, 1,124<br>(1,050-1,199) | 65, 1,060<br>(1,025-1,116) | 654, 1,179<br>(1,080-1,265) |
| **Non-AD dementia (N)** | **49** | **138** | **187** | **200** | **52** | **626** |
| NIA Amyloid stage | 14, 0 (0-0) | 58, 1 (0-2) | 116, 2 (1-2) | 117, 2 (1-3) | 35, 2 (1-2) | 340, 2 (1-2) |
| Braak-NFT stage | 15, 0 (0-I) | 66, I (I-II) | 145, II (I-III) | 165, II (I-III) | 48, III (II-III) | 439, II (I-III) |
| CERAD-NP score | 13, 0 (0-0) | 35, 0 (0-0) | 56, 0 (0-0) | 37, 0 (0-1) | 12, 0 (0-1) | 153, 0 (0-0) |
| Brain weight [gr] | 49, 926 (812-1,012) | 138, 978<br>(837-1,100) | 187, 1,036<br>(946-1,147) | 200, 1,059<br>(983-1,121) | 52, 1,029<br>(977-1,144) | 626, 1,029<br>(935-1,125) |
| **Total (N)** | **271** | **360** | **552** | **681** | **267** | **2,131** |

### 3.2 NIA Amyloid stage

The average NIA Amyloid stage was high across the AD-age-continuum, while it increased with age in the non-demented individuals (Figure 1A). In the centenarian cohort we observed that some centenarians who obtained the highest MMSE scores had accumulated amyloid up until the highest NIA Amyloid stages (Figure 1B). In contrast, some centenarians with low MMSE scores had NIA Amyloid stage of 0 (Figure 1B). Of all centenarians, 9.4% had NIA Amyloid stage 0, 35.3% had stage 1, 32.9% had stage 2, and 22.4% had stage 3 (Table 1). Upon regression of the NIA Amyloid stage to the last available MMSE score, while corrected for sex, education, and time-interval between last acquired MMSE and death, we observed that there was no association between NIA Amyloid stage and MMSE score in the centenarian cohort (β=-0.30, *P*=.60; Table 3).

**Figure 1:**
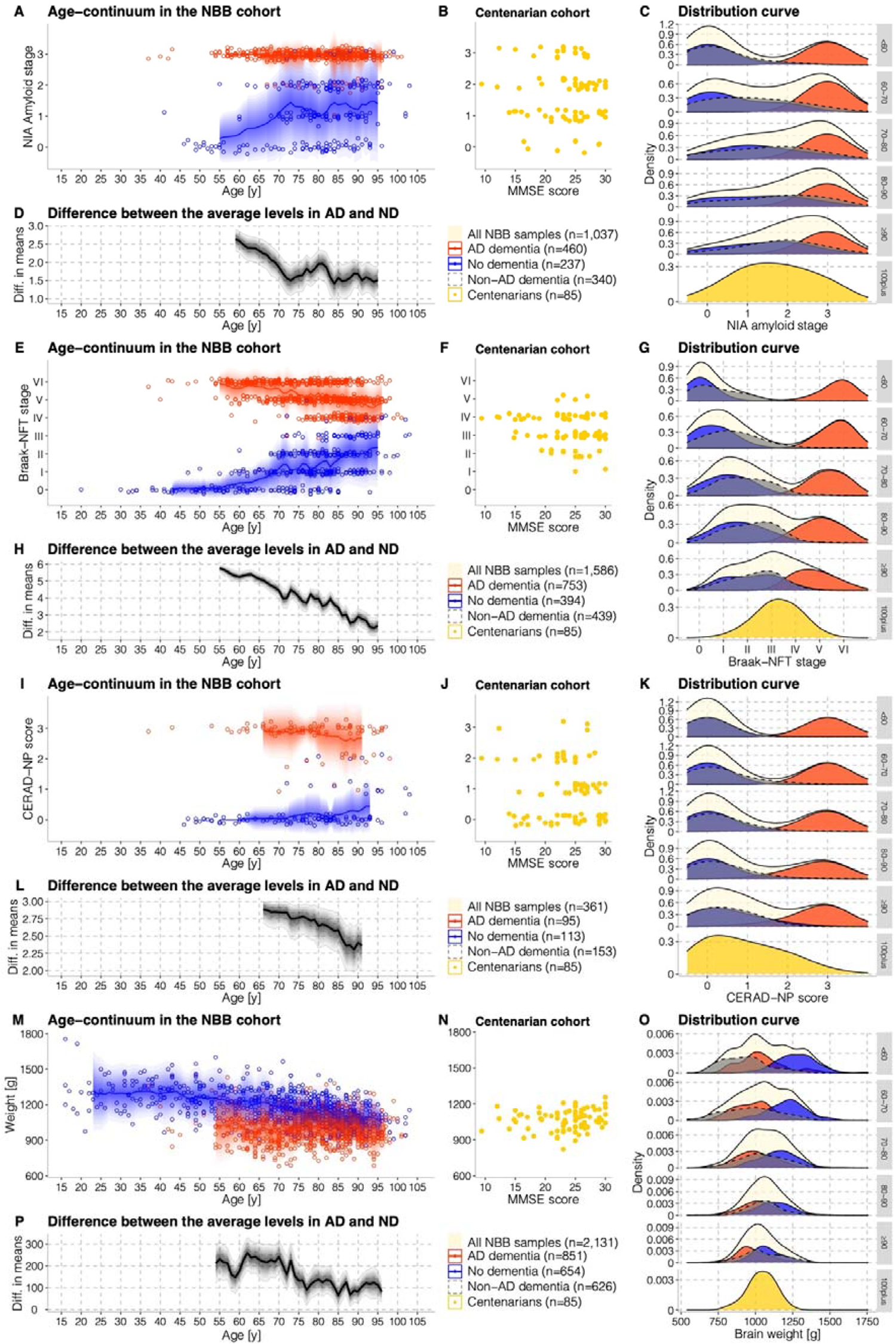
AD-associated neuropathological substrates (NIA Amyloid stage, Braak-NFT stage, and CERAD-NP score) and brain weight (corrected for sex) in the NBB and centenarian cohort. **A, E, I, M:** Mean levels of each neuropathological substrates and brain weight (corrected for sex) in AD- and ND-age-continuum in the NBB cohort ±95% confidence interval (CI). **B, F, J, H:** The levels of each neuropathological substrates and brain weight (corrected for sex) across MMSE scores in centenarian cohort. **C, G, K, O:** The distribution of the levels of each neuropathological substrate and brain weight (corrected for sex) for age intervals, i.e., ≤60, 60-70, 70-80, 80-90, ≥90, in the NBB cohort, and for the centenarian cohort. **D, H, L, P:** The difference in the average levels of each neuropathological substrates and brain weight (corrected for sex) between AD cases and non-demented controls in the NBB age-continuum ±95% CI.

**Table 3:** Associations between neuropathological substrates and MMSE score

| Neuropathology | Estimate $\beta$ (95% CI) | P value |
| --- | --- | --- |
| NIA Amyloid stage | -0.30 (-1.40, 0.81) | .60 |
| Braak-NFT stage | -1.03 (-2.21, 0.14) | .08 |
| CERAD-NP score | -0.78 (-1.87, 0.32) | .16 |
| Brain weight [gr] | 0.00 (-0.01, 0.02) | .54 |

NOTE. Using linear regression, we tested the association between the levels of each neuropathological substrate and MMSE score. The $\beta$ reflects the change in MMSE score associated with one unit increase in the level of neuropathology. The associations for AD- associated neuropathological substrates and brain weight were corrected for sex, education, and time between the last available MMSE and death.

Note that post-mortem classification of AD cases is based not only on cognitive decline, but also on having high levels of AD-associated neuropathology. Therefore, we questioned whether comparing AD and ND based on these neuropathological substrates might lead to an overestimation of the potential for each AD-associated neuropathological substrate to separate between overall decline and cognitively normal performance. Thus, we visualized the distributions of NIA Amyloid stage for age intervals ≤60, 60-70, 70-80, 80-90, ≥90 in the AD and ND cohorts, and we added a cohort of non-AD-dementia brains from the NBB, allowing a comparison with the centenarian cohort. Across all samples, we observed an age-related convergence from a bimodal distribution at younger ages to a unimodal distribution at older ages in the NBB cohort as is also observed in the centenarians (Figure 1C). This is further reflected in an age-related decrease in the difference between the average NIA Amyloid stages between AD cases and non-demented individuals, from 2.5 at age 60 to 1.5 at age ∼95 (Figure 1D).

### 3.3 Braak-NFT stage

Along the age-continuum in the NBB cohort, Braak-NFT stages ranged between 0-VI. While Braak-NFT stages were consistently higher in AD cases compared to non-demented individuals, they decreased with age in AD patients, and increased with age in non-demented individuals (Figure 1E). Braak-NFT stages in centenarians ranged between I-V (Figure 1F): none had Braak-NFT stage 0; 2.4% had stage I; 14.1% had stage II; 42.4% had stage III; 35.3% had stage IV; and 5.9% had stage V (Table 1). Braak-NFT stages did not associate with MMSE score (regression analysis: β=-1.03, *P*=.08; Table 3). Similar to NIA Amyloid stage, a convergence from a bimodal distribution of Braak-NFT stage at younger ages to a unimodal distribution in the centenarians was also observed across all samples (Figure 1G). Accordingly, the average difference between Braak-NFT stages observed in AD cases and non-demented individuals, decreased from ∼6 at age 55 to ∼2 at age 95 (Figure 1H).

### 3.4 CERAD-NP score

From age ∼75 onwards, the average CERAD-NP score increased with age in non-demented individuals and decreased in AD patients. But the increase was limited, such that the average CERAD-NP scores stayed low in non-demented older individuals and high in older AD patients (Figure 1I). In the centenarian cohort, 43.5% had CERAD-NP score 0; 29.4% had score 1; 22.4% had score 2; 4.7% (n=4) had score 3 (Figure 1J, Table 1). CERAD-NP scores did not associate with MMSE score (regression analysis: β=-0.78, *P*=.16; Table 3). We observed a stable bimodal distribution in the age-continuum across NBB samples, and a unimodal distribution was only observed in centenarian cohort, with the majority having low CERAD-NP scores (Figure 1K). In line with Figure 1I, the difference in the average CERAD-NP scores between AD and ND remained high (>2) until ages ≥90 (Figure 1L).

### 3.5 Brain weight

The brain weight of AD patients was relatively stable across age: the median female brain weight was 1,003 gr (IQR: 930-1079); the median male brain weight was 1,170 gr (IQR: 1086-1261), and included samples that weighed <750 gr. The mean sex-corrected brain weight of ND individuals at middle age was 200 gr higher than in AD cases, but decreased with 0.27% and 0.28% per year for males and females respectively (Figure 1M), until at ≥90 years the difference in average brain weights was 100 gr (Table 2, Figure 1P). None of the ND or centenarian brains had the extremely low brain weights (<750g) observed in some young demented patients (Figure 1N). A regression model indicated that brain weight was not associated with the last available MMSE score in centenarians (β=0.00, *P*=.54; Table 3). While brain weights of non-AD patients were lower than AD patients at ages <60, they united the weights of AD patients and healthy controls at higher ages (Figure 1O).

### 3.6 Correlations between AD-associated pathological scores decrease with age

Next, we merged all AD, non-AD and ND individuals from the NBB into one cohort and assessed the changes in pairwise correlations between the three pathology scores across an age-continuum (Figure 2). All pathologies were highly correlated at the youngest ages (r close to 1.0). Correlations decreased with age, in particular for the NIA Amyloid stage vs. Braak-NFT stage, which reached r=0.6 at >90 years. For the NIA Amyloid stage vs. CERAD-NP score and the Braak-NFT stage vs. CERAD-NP score, the correlation coefficients remained relatively high at r=∼0.85, at >90 years. In the centenarian cohort, the correlation coefficients for the NIA Amyloid stage vs. CERAD-NP score remained at r=∼0.75, while NIA Amyloid stage vs. Braak-NFT stage dropped to r=∼0.45. Likewise, the Braak-NFT stage vs. CERAD-NP score correlation dropped to r=∼0.55.

**Figure 2:**
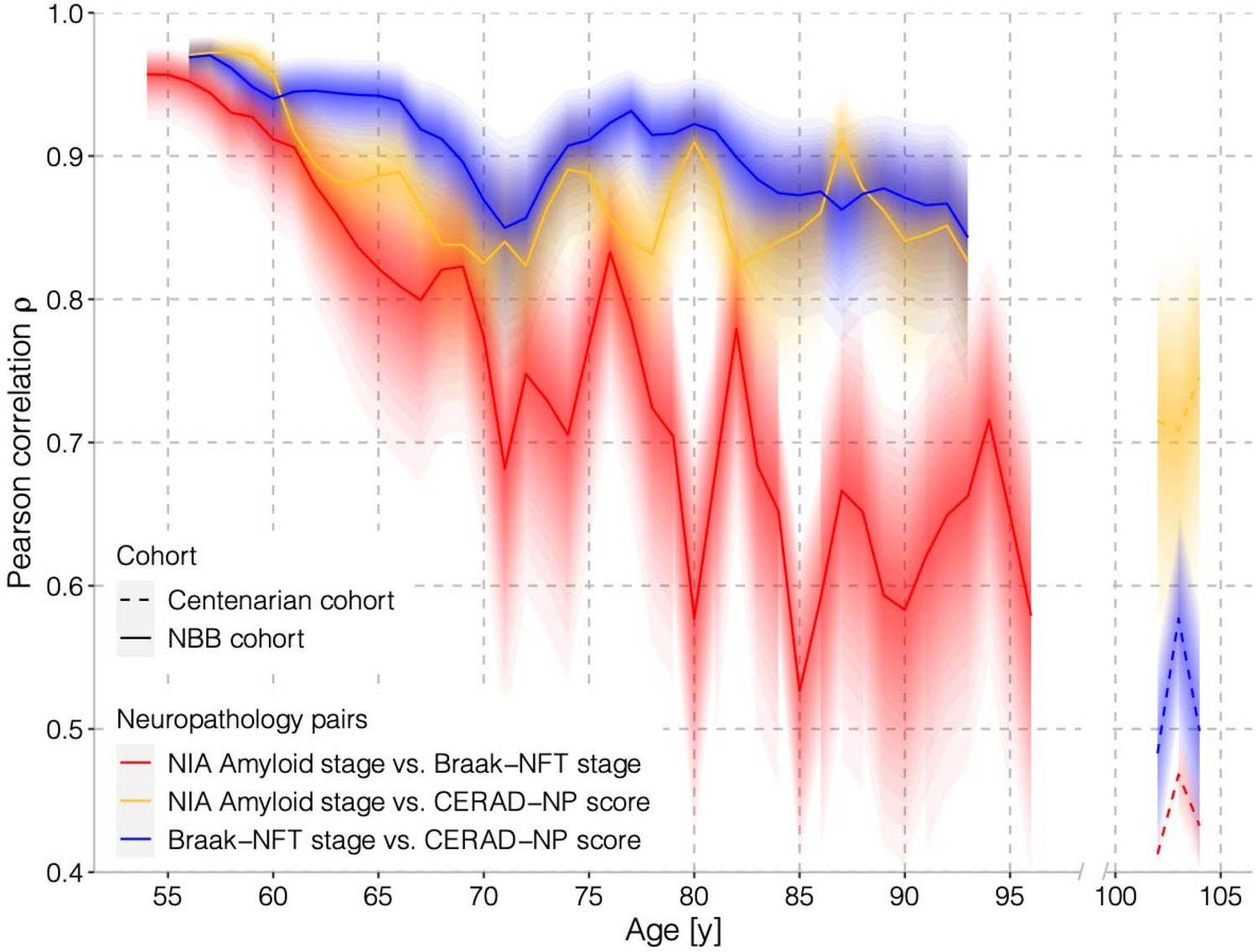
Pairwise Pearson correlation coefficient ±95% CI between AD-associated neuropathological substrates across the age-continuum in NBB and centenarian cohort separately. The NBB age-continuum includes AD patients, non-demented individuals, and non-AD demented individuals in a merged sample. The centenarian age-continuum includes all centenarians.

### 3.7 Association with demographics in the centenarian cohort

In the regression models between each of the neuropathological substrate and MMSE score, we corrected for sex, education, and time-interval between last acquired MMSE and death. We observed a significant correlation between education and MMSE (Supplementary Table S2). In contrast, there was no significant association between the levels of each neuropathological substrate and age-at-death while corrected for sex and education (Supplementary Table S3).

## 4 Discussion

In this study we observed that, with increasing age, the levels of the NIA Amyloid stage gradually increased in non-demented individuals, supporting previous reports that accumulation of amyloid is a common aspect of aging [2,3,7]. We rarely observed amyloid pathology in brains donated <65 years, while brains donated after 95 years reached amyloid levels also observed AD patients. In centenarians, NIA Amyloid stages did not correlate with cognitive performance: 9.4% of the centenarians had a NIA Amyloid stage of 0, and cognitive performance varied widely; 22.4% of the centenarians had the highest NIA Amyloid stage 3, of whom 26% had maintained high levels of cognitive performance (MMSE ≥26 [14]), suggesting that they were resilient to this pathology. A possible explanation for this may be that a considerable fraction of amyloid deposits observed in elderly, including the centenarians investigated in this study, are diffuse plaques (DPs),[8,15] which may be primarily a benign consequence of aging.

Similarly, we observed that Braak-NFT stages increased with age in non-demented individuals.[2,3,7] Most centenarians, regardless of cognitive performance, had accumulated Braak-NFT stages between I-IV; 2.4% of the centenarian brains were graded a Braak Stage I, with variable cognitive performance; 5.9% (n=5) had Braak stage V, two of whom scored 25 points and one of whom scored ≥26 points on the MMSE at last study visit, indicating resilience to the effect of NFTs. Intriguingly, the brain age-continuum indicated that Braak-NFT stages decreased with age in AD cases, which suggests that at high ages, death occurs often before the highest Braak-NFT stage is reached, presumably due to the competing risk of co-morbidity.[3,15–17]. We realized that Braak-NFT stage reflects NFT-spread in critical regions, not NFT-load; however, upon visual inspection we observed that per Braak-NFT stage, loads were not lower in centenarians compared to AD cases. Together, our results indicate that most centenarians are resilient to Braak-NFT stages ranging from I-V and/or resistant to developing higher Braak-NFT stages. In view of recent reports, we speculate that the age-related spread of NFTs may concern protective processes that remove toxic tau fibrils from the cytoplasm to store them into non- or less-toxic aggregates.[19]

With age, CERAD-NP scores also increased in cognitively healthy individuals and decreased in AD cases. However, the changes are more subtle than observed for Braak-NFT stages, such that the difference between the average CERAD-NP scores in AD cases (≥2) and non-demented individuals (≤1) remained relatively large across age. CERAD-NP scores in centenarians were mainly within the 0-2 range, and did not correlate with cognitive performance. Of the centenarians, 4.7% (n=4) had a CERAD-NP score of 3, of whom two with high cognitive performance (MMSE ≥26), suggesting that it is possible to be resilient to the highest NP scores [20].

The brain weights from non-demented individuals steadily decreased by ∼0·28% per year and approached the low brain weights observed in AD patients at ∼100 years. The brain weights of centenarians followed the age trajectory of non-demented individuals, and showed no correlation with cognitive performance. This indicates that maintaining a high brain weight is not a prerequisite for maintaining cognitive health as measured by the MMSE. After ∼40 years, loss of brain weight may be due to loss of white matter,[21] which is associated with a decrease in processing speed, which we previously observed to be characteristic for cognitive performance in the centenarian cohort.[22]

NBB distinguished between cognitive decline due to AD dementia or non-AD dementia as a consequence of AD pathology levels. The centenarian cohort included individuals with varying cognitive performance, but they were not annotated according to ND or AD or non-AD dementia. Thus, a fair comparison of the centenarian cohort with the AD and ND cohorts necessitated the addition of non-AD demented individuals. We found that, with increasing age, the distribution of the neuropathology scores of the non-AD demented individuals united the bimodal distributions of the ND and AD levels for amyloid and NFTs, but not for NPs. This suggests that amyloid and tau loads should be interpreted with caution when used for AD diagnostic purposes in the oldest old.

Notably, the reduced association between pathology substrates and cognitive performance coincides with an age-related decrease in the correlation between the levels of different neuropathological substrates. In fact, at younger ages we found an almost perfect correlation between the different substrates across the NBB sample, indicating that, a high level of one substrate correlates with having high level of another. The decrease was most apparent in the correlation between NIA Amyloid stage and Braak-NFT stage, indicating that the disease processes that lead to the buildup of these substrates in the elderly might be partly independent.[23,24] However, although lower, the correlation was still significant in the centenarian brains, which indicates that substantial dependency between the two substrates remains. Therefore, if a centenarian is resilient and/or resistant to one substrate, they may also be resilient and/or resistant to the other substrate.

There are several strengths and limitations that apply to this study. A strength is that we objectively tested cognitive performance in centenarians close to the time of brain donation, such that we have a uniquely adequate representation of brain function. The centenarians in this cohort self-reported to be cognitively healthy at study inclusion, after which they were exposed to a dementia incidence rate of ∼40% per year.[25] As the centenarian brains were donated 1-5 years after study inclusion, our analysis illuminates the neuropathological changes associated with transition from cognitive health to cognitive decline in the oldest old. However, we previously observed that average cognitive decline across the sample was mild (−1 MMSE point), and mostly driven by those who had a lower MMSE at study inclusion,[26] therefore we acknowledge that the transition to late-stage dementia remains unaddressed. A limitation of this study is the use of one measure for cognitive performance (MMSE).[27] However, earlier results of the 100-plus Study indicated that an aggregate score across all cognitive domains correlated well with the MMSE score applied on the same day,[26] suggesting that the MMSE score does have its value for interrogating overall cognitive performance. Additionally, MMSE was the first test in our testing battery, such that “MMSE at last visit” was available for almost all centenarians. Lastly, we note that the centenarians in the 100-plus cohort have a higher education compared to their birth-cohort peers,[11,26] such that cognitive reserve may contribute to their resilience against neuropathological substrates.[26] Furthermore, while there was a clear association between age and the accumulation of pathology in the age continuum, we did not identify such a relation in the centenarian cohort, suggesting that either the assessed age interval is too small to identify such a relation, or that these centenarians have unique mechanisms underlying resistance and resilience, that are not directly age-related. Technical strengths are that all brains analyzed in this study were evaluated by two consecutive neuropathologists, keeping interrater variability to a minimum.

Summarizing, the accumulation of amyloid and tau is clearly associated with disease in younger individuals, but this association decreases with age. This suggests that the age-related build-up and spread may be relatively benign in this population of centenarians. NPs retain a strong association with cognitive performance during the aging process, although we see that some centenarians are resilient to the highest levels. This observed resilience may be progressively selected for during the aging process. Increased resilience may be supported by a favorable genetic constellation, which we previously observed in the centenarians in our cohort: relative to a middle-aged population, the centenarian genomes were depleted of AD risk-alleles (including the strong risk-increasing *APOE* ε4 allele) and enriched with the genetic variants that protect against AD.[28] An alternative explanation for the observed resilience in the oldest old may be that the substrate-subtypes that build-up with age are less toxic compared to those that build-up in younger individuals, as has been shown for amyloid deposits.[8,15] However, our findings also make room for the suggestion that the accumulation of misfolded proteins at younger ages is a consequence of malfunctioning brain processes, but that the protein aggregates themselves are not necessarily toxic. It is likely that these explanations differentially contribute to the resilience observed in the oldest old for specific protein aggregates. Future studies should focus on the differentiation between pathological features associated with age related tauopathies such as PART (primary age related tauopathy) and ARTAG (aging-related tau astrogliopathy). In addition, the presence of co-pathology such as alpha-synuclein, TDP inclusions and hypoxic lesions should be considered to explain the loss of association between AD pathology and cognitive performance in the centenarians.

## Data Availability

The datasets generated during and/or analyzed during the current study are available from the corresponding author upon reasonable request.

## Acknowledgements

We thank and acknowledge all participating centenarians and their family members and thank the Netherlands Brain Bank staff for the great cooperation.

## Author contribution statement

ABG, AJMR, SR and NBB collected and performed the neuropathological characterization of the brain tissues donated to the 100-plus Study. NBB and AJMR collected and provided additional data from the Netherlands Brain Bank cohort. MZ, ABG and MH have performed the data analysis. MZ, ABG, SR, MH, JJMH and HH have written the manuscript. JJMH and HH supervised the research. PS, MJTR, MH, JJMH and HH were involved in designing the study. MZ, ABG, SR and HH have verified the underlying data. All authors read and approved the manuscript.

## Data sharing statement

Data can be made available upon request.

