## Supplementary Material for "Resilience and resistance to Alzheimer’s disease-associated neuropathological substrates in centenarians: an age-continuous perspective"

^co-last authors

†Corresponding author: Dr. Henne Holstege, Alzheimer Centrum Amsterdam, Amsterdam UMC, De Boelelaan 1118, 1081 HZ Amsterdam, The Netherlands, Tel: +31 20 4440816, Fax: +31 20 4448529,

**Conflict of interest statement**

The authors declare that they have no competing interest.

### **100-plus Study cohort**

The 100-plus Study is a prospective cohort study of centenarians who self-report to be cognitively healthy, confirmed by a proxy. The 100-plus Study was approved by the Medical Ethics Committee of the VU University Medical Center (Amsterdam, the Netherlands); procedures were conducted in accordance with the Code-of-conduct for Brain Banking and Declaration of Helsinki.[1]

### **Netherlands Brain Bank (NBB) cohort**

Procedures and information about the Netherlands brain bank (NBB) cohort can be found at https://www.brainbank.nl. Details on neuropathological diagnosis, including evaluated brain regions and staining methods, is available at https://www.brainbank.nl/brain-tissue/diagnostics/.

### **Autopsies, neuropathological assessments, and scoring strategies**

Autopsies of centenarians were performed in collaboration with the NBB. At autopsy, brain weight was measured. Methods for neuropathological characterization of post-mortem brains were described previously.[2,3] In short, Haematoxylin and Eosin (H&E) stain, Gallyas silver stain and immunohistochemistry (IHC) were performed.[4,5] Distribution of Amyloid beta peptide (Aβ) and pTau in Neurofibrillary tangles (NFTs) as well as Neuritic plaques (NPs) was determined by IHC with respectively 6F/3D and AT-8 antibodies on: frontal, motor, temporal, parietal, occipital, olfactory and entorhinal cortices; hippocampus; amygdala; nucleus caudatus and putamen (not for AT-8); nucleus accumbens, and cerebellum (only for Aβ if there were indications of a high Thal Aβ stage). Aβ distribution for centenarian cohort was evaluated according to the Thal staging (Thal Aβ phase) ranging from 0 to 5, which detects both diffuse and dense-core plaques.[6] Thal Aβ phases for centenarians were converted to **NIA Amyloid stages** following NIA guidelines as 0 or no amyloid, 1/2 or isocortical/allocortical, 3 or basal ganglia, and 4/5 or brainstem/cerebellum.[2] The brains from the NBB used in the age-continuum were donated between 1979 and 2018. Between 1991 and 2017, brains were graded for Amyloid using the Braak amyloid score (O/A/B/C).[7] After 2012, the NBB also started grading Thal Aβ phases (0 to 5).[6] To keep all the definitions consistent, we converted the Braak Amyloid stage to NIA Amyloid stages as proposed by the NIA (0-3).[2] This was done as follows: O as 0, A as 1, B as 2, and C as 3. Thal Aβ phases for NBB members were also converted to NIA Amyloid stages. NFTs and NPs were identified using a combination of Gallyas silver staining and IHC for AT-8. The temporal-spatial distribution of NFTs was evaluated according to the **Braak-NFT stages** ranging from 0 to VI.[7–9] NPs were assessed according to **CERAD-NP scores** ranging from 0 to 3.[10]

### **Supplementary tables**

**Table S1: Number of individuals for non-AD dementia subtypes**

| **Type of dementia** | **Number of individuals** | **Neuropathological diagnosis** | **Number of individuals** |
| --- | --- | --- | --- |
| Frontal Temporal Dementia | 221 | Frontal Temporal Dementia | 1 |
|  |  | Fronto-temporal dementia FUS | 10 |
|  |  | Fronto-temporal dementia Pick's disease | 35 |
|  |  | Fronto-temporal dementia TDP | 62 |
|  |  | Fronto-temporal dementia TDP + MND | 12 |
|  |  | Fronto-temporal dementia TDP type A | 15 |
|  |  | Fronto-temporal dementia TDP type B | 26 |
|  |  | Fronto-temporal dementia TDP type C | 11 |
|  |  | Fronto-temporal dementia sporadic inclusion | 2 |
|  |  | Fronto-temporal dementia tauopathy | 43 |
|  |  | Fronto-temporal dementia tauopathy PSP | 1 |
|  |  | Fronto-temporal dementia ubiquitine | 2 |
|  |  | Fronto-temporal dementia ubiquitine + MND | 1 |
| Non-AD dementia | 31 | Non-Alzheimer dementia | 24 |
|  |  | Non-classifiable senile dementia | 7 |
| Primary age-related tauopathy | 163 | Dementia + s.i.c.c./congoph. angiopathy | 2 |
|  |  | Dementia with s.i.c.c. | 78 |
|  |  | Dementia with s.i.c.c. + argyr. grains | 17 |
|  |  | Dementia with s.i.c.c. / some Lewy bodies | 66 |
| Posterior cerebral atrophy dementia | 1 | Posterior cerebral atrophy dementia | 1 |
| Parkinson's disease | 115 | Parkinson's disease with dementia | 115 |
| Vascular dementia | 95 | Dementia with cerebrovascular accident | 6 |
|  |  | Dementia with vascular encephalopathy | 16 |
|  |  | Vascular dementia | 72 |
|  |  | Vascular dementia/some Lewy bodies | 1 |
| **Total** | 626 |  | 626 |

Abbreviations: s.i.c.c., senile involutive cortical changes, also called primary age-related tauopathy (PART).

**Table S2: Summary of the results for linear regression between each neuropathological substrate and MMSE score**

|  | **Estimate β** | **Std. error** | **t-statistic** | ***P* value** |
| --- | --- | --- | --- | --- |
| **MMSE score ~** |  |  |  |  |
| **NIA Amyloid stage** | -0.30 | 0.55 | -0.53 | 0.60 |
| **+ Sex (m)** | -0.34 | 1.17 | -0.29 | 0.77 |
| **+ Education** | 0.72 | 0.32 | 2.24 | 0.03 |
| **+ Years** | -0.65 | 0.71 | -0.90 | 0.37 |
| **MMSE score ~** |  |  |  |  |
| **Braak-NFT stage** | -1.03 | 0.59 | -1.75 | 0.08 |
| **+ Sex (m)** | -0.30 | 1.15 | -0.26 | 0.80 |
| **+ Education** | 0.69 | 0.31 | 2.2 | 0.03 |
| **+ Years** | -0.44 | 0.71 | -0.62 | 0.54 |
| **MMSE score ~** |  |  |  |  |
| **CERAD-NP score** | -0.78 | 0.55 | -1.41 | 0.16 |
| **+ Sex (m)** | -0.38 | 1.15 | -0.33 | 0.74 |
| **+ Education** | 0.71 | 0.32 | 2.24 | 0.03 |
| **+ Years** | -0.63 | 0.70 | -0.89 | 0.37 |
| **MMSE score ~** |  |  |  |  |
| **Brain weight** | 0.00 | 0.01 | 0.62 | 0.54 |
| **+ Sex (m)** | -0.30 | 1.17 | -0.26 | 0.80 |
| **+ Education** | 0.65 | 0.34 | 1.93 | 0.06 |
| **+ Years** | -0.65 | 0.71 | -0.92 | 0.36 |

NOTE. Years, time between the last available MMSE and death.

**Table S3: Correlation between neuropathological substrates and age-at-death**

| **Neuropathology** | **Estimate β (95% CI)** | ***P* value** |
| --- | --- | --- |
| NIA Amyloid stage | 0.04 (-0.04, 0.13) | 0.31 |
| Braak-NFT stage | 0.03 (-0.05, 0.11) | 0.53 |
| CERAD-NP score | 0.05 (-0.04, 0.13) | 0.28 |
| Brain weight [gr] | -3.35 (-10.76, 4.06) | 0.37 |

NOTE. Using linear regression, we tested the correlation between the levels of each neuropathological substrate and age-at-death while corrected for sex and education. The β reflects the change in neuropathological levels associated with one-year increase in age.
